# Epicardial adipose tissue thickness is associated with increased COVID-19 severity and mortality

**DOI:** 10.1101/2021.03.14.21253532

**Authors:** Roopa Mehta, Omar Yaxmehen Bello-Chavolla, Leonardo Mancillas-Adame, Marcela Rodriguez-Flores, Natalia Ramírez Pedraza, Bethsabel Rodríguez Encinas, Carolina Isabel Pérez Carrión, María Isabel Jasso Ávila, Jorge Carlos Valladares-García, Pablo Esteban Vanegas-Cedillo, Diana Hernández Juárez, Arsenio Vargas-Vázquez, Neftali Eduardo Antonio-Villa, Monica Chapa-Ibarguengoitia, Paloma Almeda-Valdés, Daniel Elias-Lopez, Arturo Galindo-Fraga, Alfonso Gulias-Herrero, Alfredo Ponce de Leon, José Sifuentes-Osornio, Carlos A. Aguilar-Salinas

## Abstract

**BACKGROUND:** Increased adiposity and visceral obesity have been linked to adverse COVID-19 outcomes. The amount of epicardial adipose tissue (EAT) may have relevant implications given its proximity to the heart and lungs. Here, we explored the role of EAT in increasing the risk for COVID-19 adverse outcomes.

**METHODS:** We included 748 patients with COVID-19 attending a reference center in Mexico City. EAT thickness, sub-thoracic and extra-pericardial fat were measured using thoracic CT scans. We explored the association of each thoracic adipose tissue compartment with COVID-19 mortality and severe COVID-19 (defined as mortality and need for invasive mechanical ventilation), according to the presence or absence of obesity. Mediation analyses evaluated the role of EAT in facilitating the effect of age, body mass index and cardiac troponin levels with COVID-19 outcomes.

**RESULTS:** EAT thickness was associated with increased risk of COVID-19 mortality (HR 1.18, 95%CI 1.01-1.39) independent of age, gender, comorbid conditions and BMI. Increased EAT was associated with lower SpO2 and PaFi index and higher levels of cardiac troponins, D-dimer, fibrinogen, C-reactive protein, and 4C severity score, independent of obesity. EAT mediated 13.1% (95%CI 3.67-28.0%) and 5.1% (95%CI 0.19-14.0%) of the effect of age and 19.4% (95%CI 4.67-63.0%) and 12.8% (95%CI 0.03-46.0%) of the effect of BMI on requirement for intubation and mortality, respectively. EAT also mediated the effect of increased cardiac troponins on myocardial infarction during COVID-19.

**CONCLUSION:** EAT is an independent risk factor for severe COVID-19 and mortality independent of obesity. EAT partly mediates the effect of age and BMI and increased cardiac troponins on adverse COVID-19 outcomes.

## INTRODUCTION

Obesity is a major risk factor for disease severity related to severe acute coronavirus 2 (SARS-CoV-2) infection^1^. Patients with obesity have a higher risk for, hospitalization, intensive care unit (ICU) admission, invasive mechanical ventilation (IMV) and death ^1,2^. Obesity is associated with a state of low-grade inflammation, which in the setting of coronavirus disease (COVID-19) can result in further inflammation, endothelial dysfunction and a subsequent cytokine storm, increasing the risk for severe disease ^2–4^. In Mexico, obesity is highly prevalent; in particular, the accumulation of abdominal visceral adipose tissue (VAT) has been shown to influence COVID-19 outcomes, independent of additional comorbid conditions and risk factors.^5-8^

Visceral obesity is associated with increased cardiometabolic burden, including risk of arterial hypertension, type 2 diabetes mellitus and cardiovascular disease^5^. Accumulation of VAT results in adipose tissue dysfunction, morphological changes in adipocytes, fibrosis, altered secretion of adipokines and increased inflammation^6^. VAT has proven to be related to increased risk for severe COVID-19 and need for intensive care.^11, 12^ The engagement of SARS-CoV-2 with angiotensin converting enzyme 2 (ACE 2) in visceral fat is thought to impair the function of this enzyme, resulting in increased production of angiotensin II, thus enhancing the production of inflammatory cytokines.^11^ Levels of ACE 2 mRNA in visceral fat correlate with body mass index; this is not the case for subcutaneous fat. Epicardial adipose tissue (EAT) is an ectopic fat deposit (visceral fat), located between the myocardium and the visceral layer of pericardium, and is thought to contribute to adverse cardiovascular outcomes^7^. EAT may play a significant role in mediating the effects of obesity on COVID-19 outcomes due to its thoracic localization and may influence the cardiovascular complications of COVID-19^8–10^. In this study, we examined the association of EAT with adverse COVID-19 outcomes and explored potential pathways which may link the accumulation of EAT was mediators of well-established risk factors related to severe COVID-19.

## METHODS

### Study population

This study included consecutive patients evaluated at the Instituto Nacional de Ciencias Médicas y Nutrición Salvador Zubirán (INCMNSZ), a COVID-19 reference centre in Mexico City between 17th March and 31st May 2020 (**Supplementary Material**). Subjects were initially assessed at triage and required either ambulatory or in-hospital care for COVID-19, confirmed with computerized tomography (CT) and/or via RT-qPCR test from nasopharyngeal swabs. All patients had moderate to severe disease as defined by National Institute of Health criteria. *Moderate* illness was defined as evidence of lower respiratory disease during clinical assessment or imaging and who have saturation of oxygen (SpO_2_) ≥94% on air, and s*evere illness as* SpO_2_ <94% on air, a ratio of arterial partial pressure of oxygen to fraction of inspired oxygen (PaO_2_/FiO_2_) <300 mm Hg, respiratory frequency >30 breaths/min, or lung infiltrates >50%). All patients underwent a chest CT, and a radiologist determined the degree of pulmonary parenchymal disease and assessed epicardial fat thickness. In addition, a medical history, anthropometric measurements and laboratory tests were obtained. The electronic files of each patient were retrospectively reviewed to document the outcomes during hospitalization. All proceedings were approved by the research and ethics committee of the INCMNZ (Ref. 3383) and informed consent was waived due to the nature of the study.

### Laboratory and clinical measurements

Clinical variables and laboratory measures were obtained at the time of initial evaluation. Physical examination included: weight, height, body mass index (BMI, calculated as weight in kilograms divided by squared height in meters), pulse oximeter saturation (SpO2), respiratory rate (RR), temperature and arterial blood pressure (BP). Laboratory measurements included: full blood count, chemistry panel including liver function tests, C-reactive protein (CRP), fibrinogen, D-dimer, ferritin, troponin I (TPNI), erythrocyte sedimentation rate (ESR) and procalcitonin levels. The blood samples were processed at the central laboratory of the INCMNSZ.

### Epicardial adipose tissue measurements

All patients underwent unenhanced CT scans including low-dose CT and two ultra-low-dose CT protocols and commonly reported imaging features of COVID-19 pneumonia were captured. Thorax CT was performed using a 64-slice scanner (GE MEDICAL SYSTEMS Revolution EVO). Epicardial adipose tissue (EAT), the adipose tissue located between the visceral layer of the pericardium and the surface of the heart was measured. EAT thickness was measured at 3 points (right atrioventricular fossa, left atrioventricular fossa, and anterior interventricular fossa) in the reformatted 4-chamber view using the multiplanar reconstruction (MPR) tool on the workstation (19, 20, 21). The maximum thickness of the EAT was recorded, from the surface of the myocardium to the pericardium (measured perpendicular to the pericardium). The measurements were made on 2 different occasions, obtaining a total of 6 measurements; the average of them was used in the statistical analysis. Extrapericardial adipose tissue (PAT) was quantified with the volume measurement tool with the Carestream system of the workstation. The thickness of the thoracic subcutaneous adipose tissue (TscAT) was measured from the anterior border of the sternum to the skin, at the level of the mitral valve in the axial plane of the tomography as a surrogate for subcutaneous adipose tissue. The 80^th^ gender-specific percentile of EAT thickness was obtained and used as the threshold to define increased EAT thickness, given the unavailability of population-based percentiles. This was used in categorical analysis, which are supplementary to the analysis of continuous variables. In addition, chest CT findings were recorded and used to evaluate severity of COVID infection.

### COVID-19 outcomes

Outcomes included mortality and a composite of death and requirement for invasive mechanical ventilation, which was used to define severe COVID-19 given the potential of unobserved severe cases in a setting of limited healthcare facilities. For time-to-event analyses, time from self-reported symptom onset prior to evaluation until last follow-up (censoring) or death, whichever occurred first was estimated. The COVID-19 specific 4C mortality score was estimated; this reflects increased risk of mortality due to COVID-19^11^.

### Statistical analyses

The clinical characteristics of subjects with visceral obesity (EAT > 80^th^ percentile) were compared according to presence or absence of obesity, as assessed by BMI, using Student’s t-test or Mann-Whitney U according to variable distribution for continuous variables and using chi-squared tests for categorical variables. To ensure variable symmetry prior to modeling, the *bestNormalize* R package was used and identified the better transformation as out-of-sample normalization via 10-fold cross-validation with 5 repeats for all CT-derived measurements. All statistical analyses were conducted using R 4.0.2.

#### Assessment of risk based on thoracic fat distribution

The CT-derived adipose tissue measures and body-mass index (BMI) were compared between cases who eventually required mechanical ventilation or died during hospitalization and those who did not, using Student’s t-test. Linear regression analysis was used to assess which clinical features were independently associated with EAT thickness in COVID-19 patients. To assess the predictive ability of each thoracic adipose tissue segment, Cox proportional risk regression models were generated including an unadjusted model (Model 1) and models adjusted for age, gender and number of comorbid conditions (Model 2) and further adjustments by BMI to assess independent risk after adjustment (Model 3). In addition, the construct of increased EAT using the definition of >80^th^ gender-specific percentile of EAT thickness was assessed using the same models. To assess the impact of thoracic adipose tissue compartments on the risk of severe COVID-19, similar models were adjusted using fixed effects logistic regression models. To assess the independent effect of BMI, Models 1 and 2 were fitted for normalized BMI values for both outcomes. Finally, we included all thoracic adipose tissue depots in a multivariable Cox regression model to assess independency for the risk for severe COVID-19 attributable to EAT, adjusted for sex, age, BMI and comorbidities.

#### Post-estimation simulation of mortality risk

The 1-unit incremental risk associated with normalized EAT measures was evaluated using Cox regression models adjusted for age, gender, number of comorbid conditions and BMI-This was further stratified by BMI-determined obesity status to evaluate the independent effect of EAT thickness in predicting mortality risk related to COVID-19 using the *simPH* and *survival* R packages.

#### Mediation analyses

The first hypothesis was that EAT was partially mediating the mortality risk related to BMI, given the association of increases in visceral and epicardial adipose tissue with mortality risk^12^. The secondary hypothesis was that EAT may induce cardiac damage and that this could increase COVID-19 severity and mortality risk^8^. Finally, we evaluated whether EAT may induce a hypercoagulable state via D-dimer or fibrinogen which may contribute to severe COVID-19 or death. To test these hypotheses, model-based casual mediation analyses were conducted using the *mediation* R package, adjusting logistic regression models by age, gender, number of comorbid conditions or BMI as required. Confidence intervals were estimated using White’s heteroskedasticity-consistent estimator for the covariance matrix, derived from quasi-Bayesian Monte Carlo simulation based on normal approximation.

## RESULTS

### Study population

We included 748 patients with confirmed COVID-19, with a mean age of 51.22±13.62 years, male predominance (n=470, 63.1%), and a mean BMI of 30.25±5.71. Median follow-up was 14.0 days (IQR 10.0-20.0) and 629 patients required hospitalization (85.5%). Overall, 138 patients received invasive mechanical ventilation (18.5%),164 in-hospital deaths (21.9%) were recorded and 236 cases of severe COVID-19 were documented (31.6%, **Supplementary Material**). Type 2 diabetes (T2D) was present in 191 patients (26.3%), 300 patients had obesity (44.4%) and 282 were overweight (41.8%). Median EAT thickness was 9.67mm (IQR 7.67-12.0), median TscAT was 16mm (IQR 11.0-23.0) and PAT was present in 451 patients (60.5%).

### Thoracic fat distribution and COVID-19 severity

Cases with severe COVID-19 had greater EAT thickness, and BMI with no significant difference in PAT and lesser TscAT compared to non-severe COVID-19. In fatal cases, EAT thickness and PAT volume were significantly higher; BMI was greater but failed to reach statistical significance and TscAT showed no difference between fatal and non-fatal cases (**Figure 1**). When stratifying cases according to BMI-defined obesity, those who had increased EAT were older, had worsened respiratory parameters, altered coagulation and overt inflammation, and had higher rates of severe outcomes and intubation as well as higher mortality scores independent of BMI status (**Table 1**). EAT thickness was independently associated with male gender (β=-0.236, 95%CI -0.382, -0.090), oxygen saturation (β=-0.014, 95%CI -0.019,-0.008) and higher 4C mortality scores (β=0.063, 95%CI 0.039, 0.087, adjusted R^2^=0.103). In a second model, EAT thickness was associated with increased levels of ultrasensitive cardiac troponins (β=0.0002, 95%CI 0.00005, 0.0004), BMI (β=0.034, 95%CI 0.021, 0.048) and age (β=0.017, 95%CI 0.011, 0.023, adjusted R^2^=0.081), even after adjustments for C-reactive protein levels.

**Table 1.** Comparison of demographic and clinical markers of COVID-19 patients comparing epicardial adipose tissue thickness (EAT) (increased >80^th^ sex-specific percentile or not) stratified by obesity status as defined by body-mass index (BMI).

| Parameter | Overall | Non-Obese |  |  | Obese |  |  |
| --- | --- | --- | --- | --- | --- | --- | --- |
|  |  | EAT not-increased<br>(n=368) | EAT increased<br>(n=74) | P-value | EAT not-increased<br>(n=230) | EAT increased<br>(n=76) | P-value |
| Age (years) | 51 (41-61) | 51.5 (13.7) | 56.9 (15.2) | 0.006 | 48.4 (12.9) | 52.5 (13.8) | 0.002 |
| Male Sex (%) | 470 (63.1) | 227 (62.2) | 49 (66.2) | 0.602 | 149 (64.7) | 45 (59.2) | 0.944 |
| Low-Socioeconomic Status (%) | 530 (71.2) | 262 (71.9) | 53 (71.6) | 0.682 | 161 (71.8) | 54 (71.1) | 0.682 |
| Severe outcome (%) | 236 (31.5) | 91 (24.7) | 34 (45.9) | 0.003 | 78 (33.9) | 33 (43.5) | 0.025 |
| Intubation (%) | 138 (18.4) | 46 (12.5) | 19 (12.5) | 0.006 | 54 (23.4) | 19 (25) | 0.002 |
| Mortality (%) | 164 (21.9) | 67 (18.2) | 22 (29.8) | 0.036 | 48 (20.8) | 27 (35.5) | 0.182 |
| Arterial Hypertension (%) | 212 (29.2) | 88 (24.9) | 23 (31.9) | 0.277 | 73 (32.2) | 28 (37.3) | 0.039 |
| Type 2 diabetes (%) | 191 (26.3) | 87 (24.6) | 23 (32.3) | 0.226 | 55 (24.2) | 26 (34.7) | 0.857 |
| Time since DM2 diagnosis (years) | 7 (3-14.8) | 8 (5-14.5) | 6 (2-12.5) | 0.370 | 6 (1-15) | 6 (5-10) | 0.773 |
| Smoking status (%) | 47 (7.7) | 21 (7.26) | 6 (7.25) | 0.792 | 16 (8.2) | 4 (6.2) | 0.826 |
| CKD (%) | 26 (3.6) | 14 (3.2) | 4 (5.5) | 0.780 | 3 (1.3) | 5 (6.7) | 0.345 |
| CVD (%) | 19 (2.6) | 9 (2.6) | 1 (1.4) | 0.868 | 6 (2.6) | 3 (4.0) | 0.774 |
| Cirrhosis (%) | 6 (0.82) | 2 (0.05) | 1 (1.4) | 0.999 | 2 (0.8) | 1 (1.3) | 0.998 |
| BMI (kg/m2) | 30.4 (11.2) | 27 (25.1-28.7) | 27.5 (25-29) | 0.061 | 33.2 (31-37) | 34.1 (31-36) | 0.013 |
| Respiratory Rate (rpm) | 28 (11.2) | 25 (20-30) | 30 (26-36) | <0.001 | 26 (22-33) | 28 (24-39) | <0.001 |
| Heart Rate (bpm) | 101 (18.6) | 102 (90-113) | 103 (90-115) | 0.529 | 102 (90-115) | 107 (90-115) | 0.385 |
| Systolic Arterial Pressure (mmHg) | 121 (110-132) | 120 (110-130) | 120 (110-130) | 0.587 | 127 (113-135) | 128 (111-134) | 0.771 |
| Diastolic Arterial Pressure (mmHg) | 75 (69-80) | 73 (69-80) | 70 (66-80) | 0.397 | 78 (70-82) | 76 (68-80) | 0.343 |
| Oxygen saturation (%) | 86 (77-89) | 88 (80-90) | 79 (63-87) | <0.001 | 85 (79-89) | 82 (60-87) | <0.001 |
| C-reactive protein | 14.3 (7.1-22.4) | 14.93 (9.02-23.20) | 19.3 (10.8-28.5) | <0.001 | 14.4 (7.9-21.2) | 19 (9-28.6) | <0.0001 |

**Figure 1.**
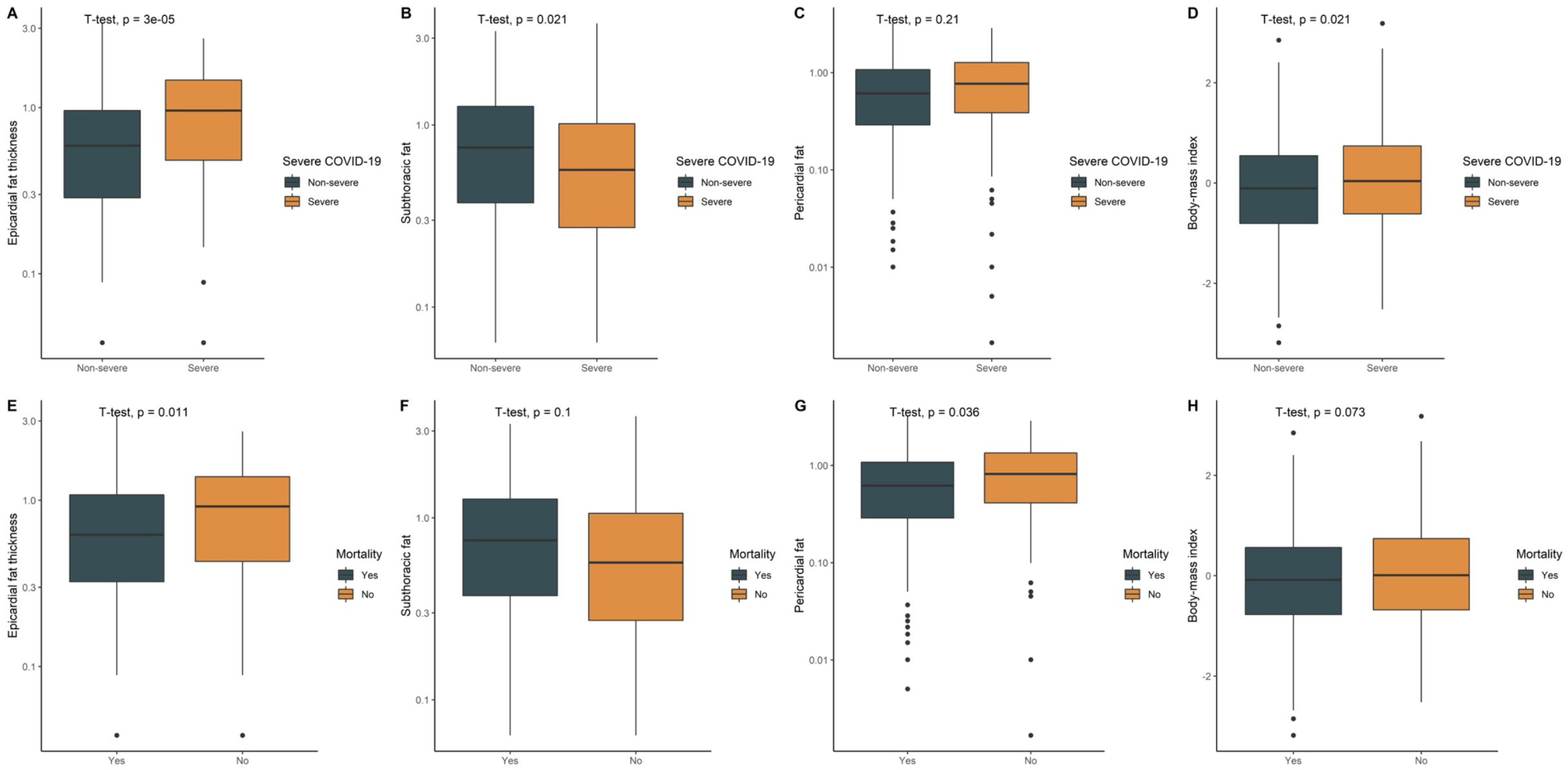
Boxplots comparing transformed levels of epicardial fat thickness, pericardial fat, subthoracic fat and body-mass index (BMI), according to severe vs. non-severe COVID-19 (IVM, A-D) or COVID-19 mortality (E-H).

#### EAT thickness is an independent predictor of COVID-19 mortality

Using Cox regression analysis, EAT thickness was a predictor of COVID-19 mortality and severe COVID-19, even after adjustment for age, gender, number of comorbid conditions and BMI (**Tables 2-3**). Using post-estimation simulation, we observed monotonic increases in mortality risk with increasing normalized EAT thickness both in obese and non-obese subjects (**Figure 2**). Subjects who had increased EAT had higher risk of both mortality and risk for severe COVID-19. Similarly, after assessing the independent effect of other adipose tissue segments, PAT volume was associated with increased mortality in univariate analyses but failed to retain significance after adjustments for age, gender, number of comorbid conditions and BMI. Similarly, TscAT thickness was associated with increased risk of mortality and severe COVID-19 after adjustment for age, gender, and number of comorbid conditions but after adjustment for BMI this association failed to maintain statistical significance. In contrast, BMI was associated with both mortality and severe COVID-19 even after adjustment for covariates (**Tables 2-3**).

**Table 2.** Cox proportional risk regression models to predict mortality related to COVID-19 using CT-scan derived fat measures and BMI transformed using repeated out-of sample 10-fold cross-validation and the definition of visceral obesity as epicardial fat thickness values >80th sex-adjusted percentiles for the population. Model 1: Univariate, Model 2: Ajusted for age, gender and comorbid conditions, Model 3: Adjusted for age, gender, comorbid conditions and BMI

| Model | Model 1 |  | Model 2 |  | Model 3 |  |
| --- | --- | --- | --- | --- | --- | --- |
|  | HR (95%CI) | P-value | HR (95%CI) | P-value | HR (95%CI) | P-value |
| Epicardial fat thickness | 1.290 (1.108-1.501) | 0.001 | 1.212 (1.033-1.423) | 0.019 | 1.180 (1.003-1.389) | 0.047 |
| Extrapericardial fat volume | 1.223 (1.036-1.445) | 0.018 | 1.108 (0.924-1.328) | 0.269 | 1.049 (0.870-1.265) | 0.616 |
| Subthoracic fat volume | 1.067 (0.907-1.256) | 0.434 | 1.230 (1.019-1.483) | 0.031 | 1.117 (0.889-1.404) | 0.343 |
| Body-mass index | 1.091 (0.931-1.280) | 0.283 | 1.262 (1.042-1.529) | 0.018 | - | - |
| Increased EAT (>p80, sex-based ) | 1.570 (1.123-2.196) | 0.008 | 1.433 (1.023-2.009) | 0.037 | 1.409 (1.006-1.975) | 0.046 |

**Table 3.**
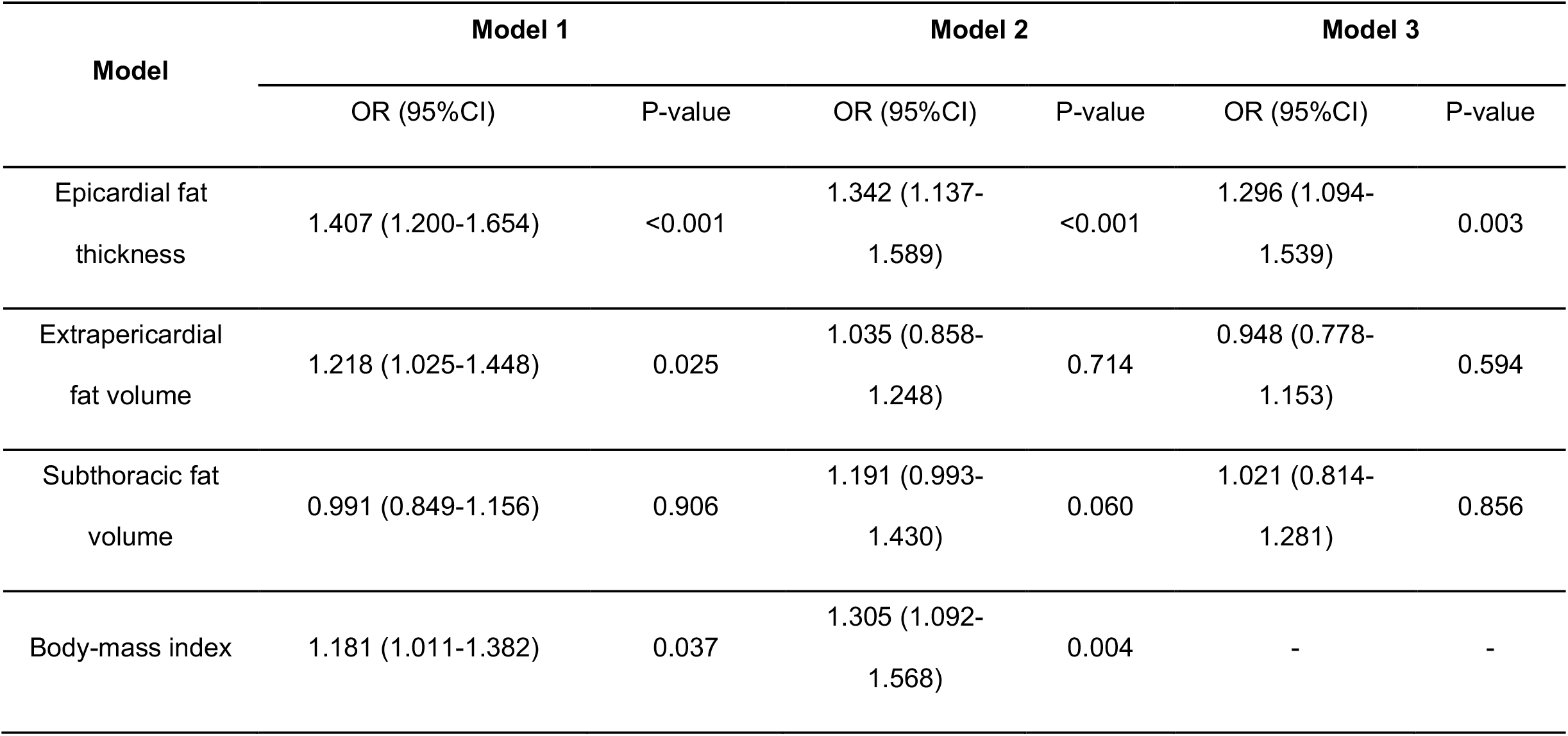
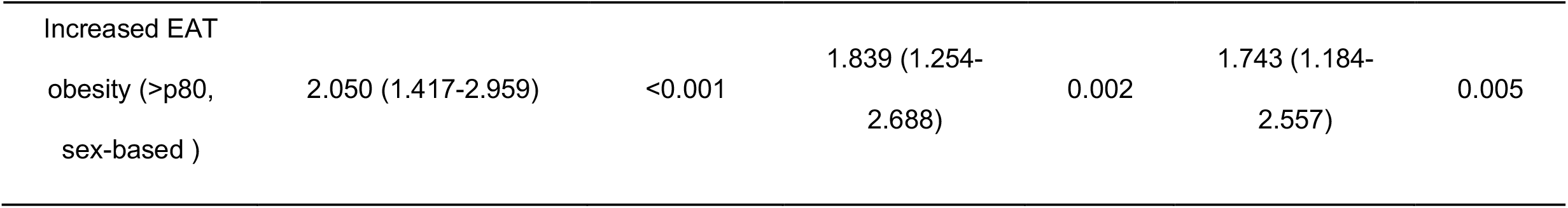
Logistic regression models to predict severe COVID-19, using CT-scan derived fat measures and BMI transformed with repeated out-of sample 10-fold cross-validation and the definition of increased epicardial fat thickness (EAT) values >80th sex-adjusted percentiles for the population. Model 1: Univariate, Model 2: Adjusted for age, gender and comorbid conditions, Model 3: Adjusted for age, gender, comorbid conditions and BMI.

**Figure 2.**
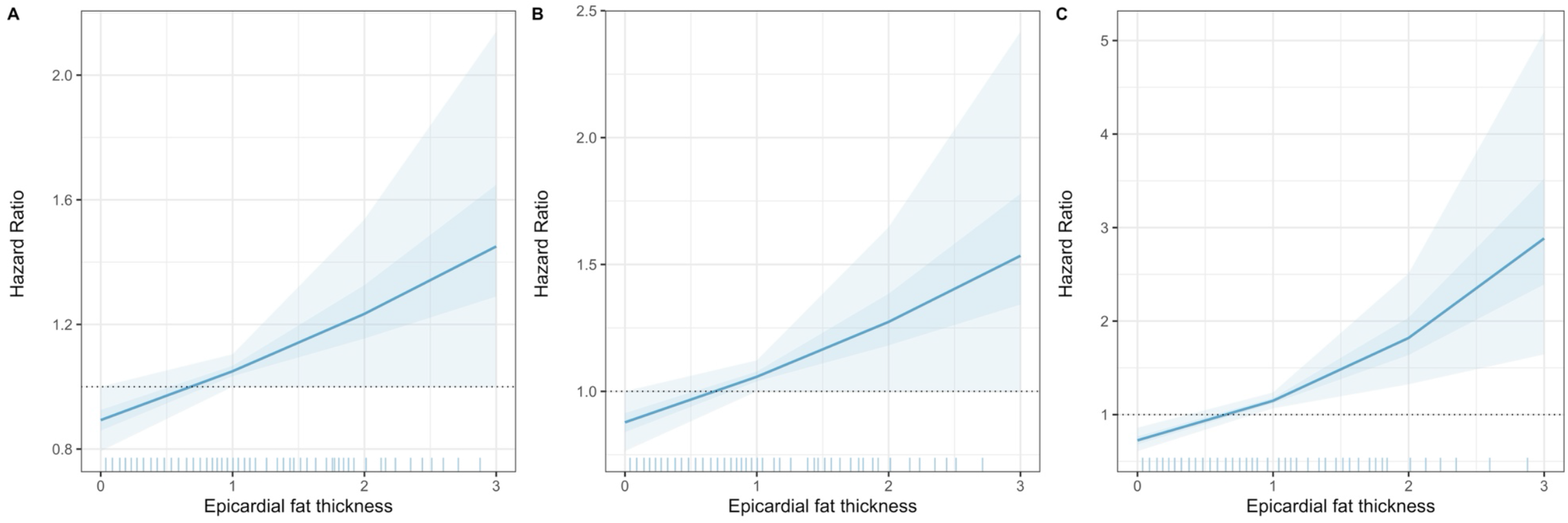
Post-estimation simulation of the risk of epicardial fat thickness transformed by repeated out-of sample 10-fold cross-validation adjusted for age, gender, comorbid conditions and BMI (A) stratified in non-obese (B) and obese (C) cases with confirmed COVID-19.

In multivariable analyses for mortality, increased EAT was the only compartment which independently showed increased risk of mortality (HR 1.26, 95%CI 1.07-1.48), with no significant risk attributable to TscAT (HR 1.08, 95%CI 0.85-1.35) or PAT (HR 1.12, 95%CI 0.93-1.34) after adjustment for sex, BMI, age and comorbidities. For severe COVID-19, we similarly observed increased risk for EAT (OR 1.33, 95%CI 1.12-1.59) but not for TscAT (OR 0.98, 95%CI 0.77-1.25) or PAT (OR 0.87, 95%CI 0.72-1.09), after adjustment for similar confounders. This confirms an independent contribution of EAT compared to other thoracic adipose tissue compartments for adverse COVID-19 outcomes.

#### EAT partially mediates the effect of BMI on COVID-19 mortality

Given the association of BMI, and increased cardiac troponins on COVID-19 mortality, severe COVID-19 and EAT thickness, two causal mediation models were explored. We explored EAT thickness as a mediator of BMI and in increasing risk of COVID-19 mortality and severe COVID-19 (**Figure 2**). EAT thickness mediates 19.4% (95%CI 4.67-63.0%) of the effect of BMI on the risk of severe COVID-19, and 12.8% (95%CI 0.03-46.0%) of the effect of BMI on risk of COVID-19 mortality, after adjustment for age, gender and number of comorbid conditions (**Table 4**).

**Table 4.**
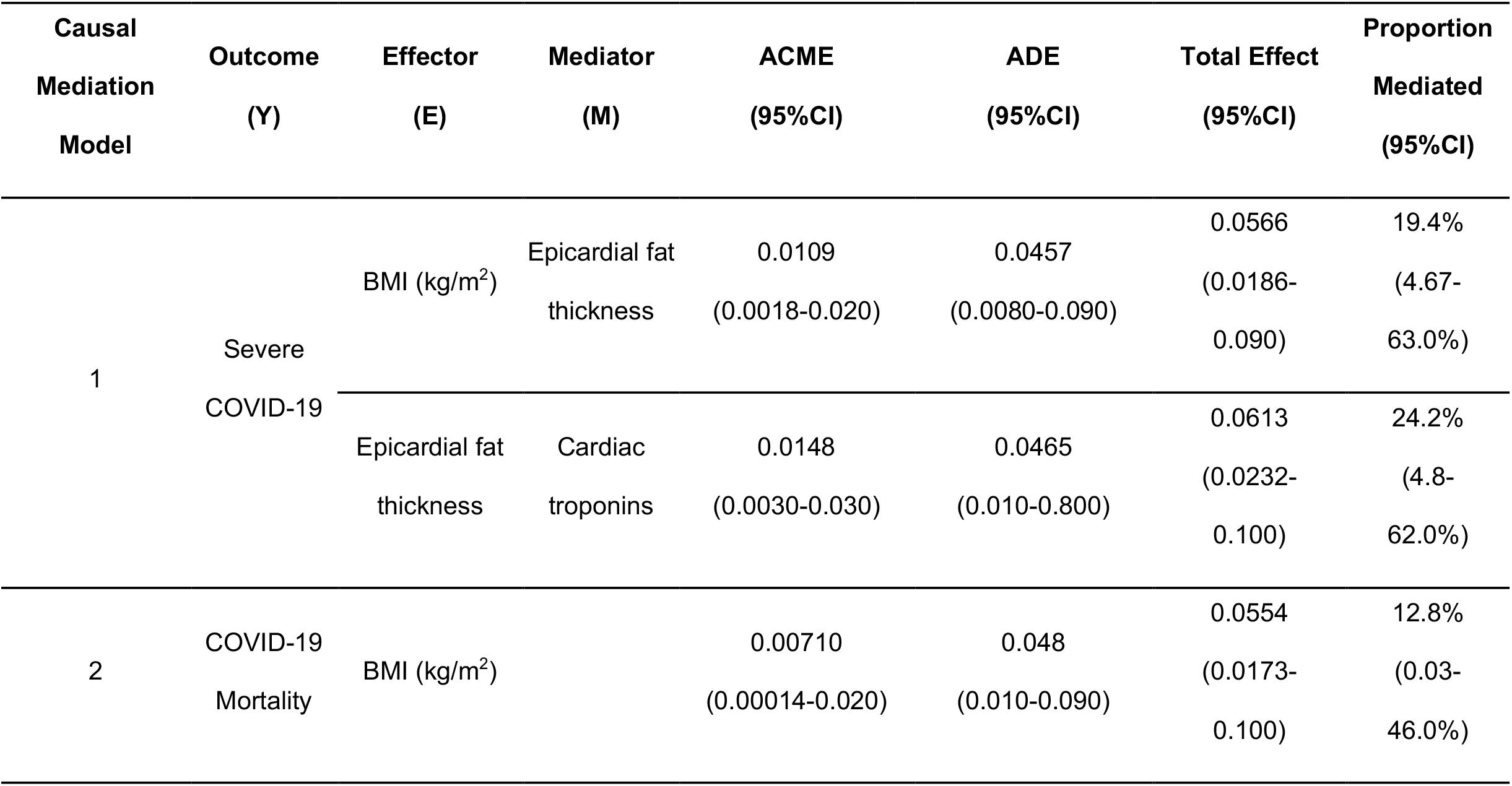
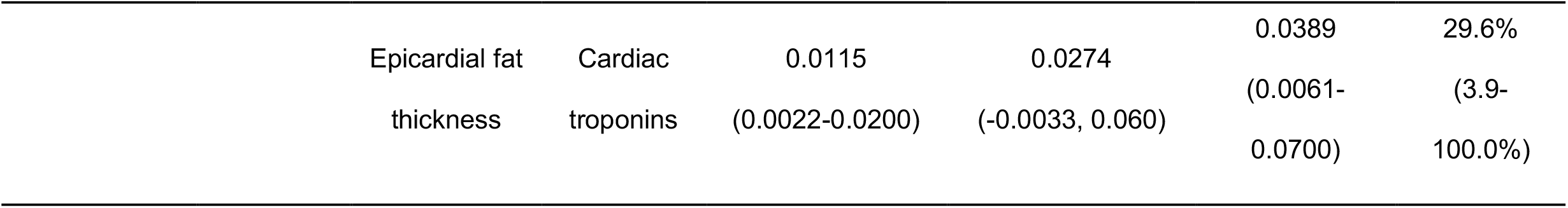
Causal mediation models. Firstly, evaluating the effect of age and BMI, on COVID-19 outcomes via epicardial fat thickness. Secondly, the role of epicardial adipose tissue thickness on COVID-19 outcomes via cardiac troponins. Abbreviation: ACME: average causal mediation effects; ADE: average direct effects

#### EAT-induced heart damage mediates COVID-19 adverse outcomes

The second causal mediation model explored whether increased risk for COVID-19 outcomes was partially mediated by EAT-induced heart damage (**Figure 3**). Cardiac troponins mediate 24.2% (95%CI 4.8-62.0%) of the effect of EAT on the risk of severe COVID-19 and 29.6% (95%CI 3.9-100.0%) of the effect of EAT on the risk of COVID-19 mortality adjusted for age and gender. Finally, we identified that EAT may mediate risk of death in COVID-19 via increased fibrinogen levels by about 15.2% (95%CI 3.8-47.0%), and risk for severe COVID-19 by about 13.5% (95%CI 4.0-29.0%), adjusted for age, sex, BMI and comorbidities. Regarding cardiovascular events, there were 19 registered myocardial infarctions, of whom 13 died. EAT thickness is a significant predictor of myocardial infarction in COVID-19 (OR 1.78, 95%CI 1.09-2.97); increased EAT thickness increased risk for myocardial infarction in COVID-19 (OR 2.83, 95%CI 1.07-7.31), after adjustment for age, gender and comorbid conditions

**Figure 3.**
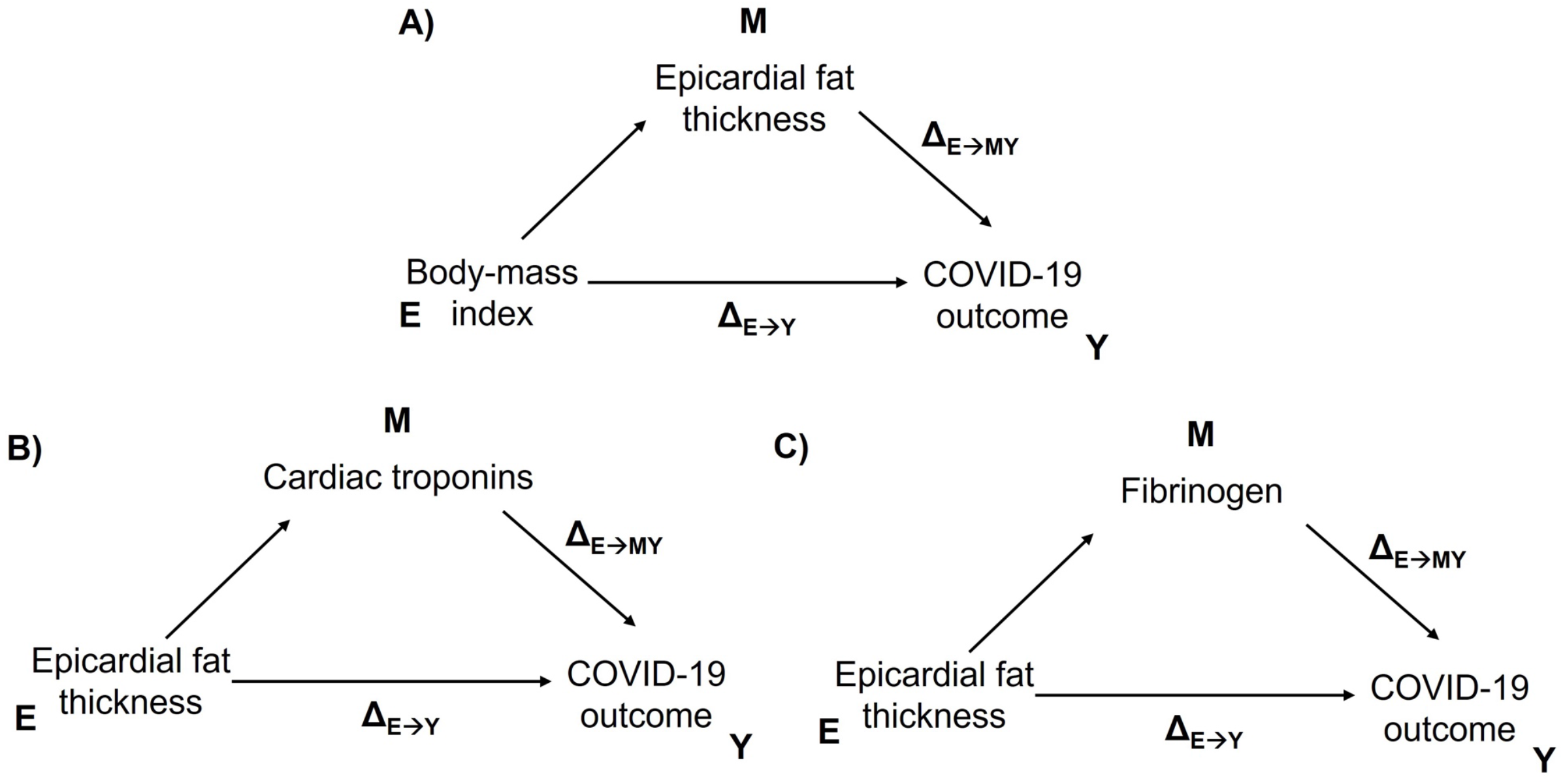
Diagrams of causal mediation models evaluating the role of epicardial adipose tissue (EAT) thickness as a mediating factor of the impact of body-mass index (BMI) on severe COVID-19 and mortality (COVID-19 outcome). Secondary causal mediation models were developed to investigate the association of ultrasensitive cardiac troponins and fibrinogen as mediators of the relationship between EAT and adverse COVID-19 outcomes via direct heart damage or a pro-coagulant state.

## DISCUSSION

In this study, EAT thickness shows a significant association with COVID-19 severity and mortality independent of BMI, age, and comorbid conditions. Subjects with increased EAT had higher levels of COVID-19 severity markers independent of BMI. Overall, EAT thickness was associated with decreased oxygen saturation, male gender, and increased COVID-19 severity, and showed an independent association with increased ultrasensitive cardiac troponin levels. Furthermore, EAT thickness partly mediated the effect of BMI related risk on COVID-19 adverse outcomes and may lead to a pro-coagulant state with myocardial damage in the setting of severe COVID-19.

The effect of EAT on risk of COVID-19 mortality and severe COVID-19 was independent of the other thoracic adipose tissue compartments. TscAT and PAT did not show an increased risk for adverse outcomes or mortality independent of BMI. This finding is probably explained by the unique characteristics of each thoracic fat compartment. Extra-pericardial adipose tissue (PAT) is located on the outer surface of the fibrous pericardium, it differs from epicardial adipose tissue (EAT) in its embryonic origin and blood supply^13^. It receives nourishment from the pericardiophrenic artery, while EAT has the same embryonic origin as visceral abdominal fat and is supplied by the coronary arteries. Presence of PAT is associated with obesity-related diseases similar to TscAT. Researchers evaluating pericardial fat (this includes both extrapericardial fat and epicardial fat) have reported an independent association with risk of cardiovascular disease^14^. This is in line with our results, confirming that adipose tissue distribution, in particular the presence of visceral adiposity more so than total adipose tissue, may play a significant role in modulating the adaptive responses to SARS-CoV-2 infection, and thus increase disease severity in COVID-19^15^.

The metabolic and inflammatory role of VAT and its role in cardiovascular and metabolic health is well documented^16^. Beyond the risk associated with whole-body adipose tissue as assessed by BMI, VAT depots significantly contribute to the increased cardiovascular risk profile and low-grade pro-inflammatory state observed in patients with obesity^17^. Increased accumulation of VAT has been associated with increased deposition of ectopic fat, particularly in the liver and in the epicardium as EAT^18^. Notably, EAT shares microcirculation with the myocardium, favoring the rapid exchange of metabolic and inflammatory products^19^; the implication of this anatomical relationship suggests that, under acute or chronic inflammatory conditions, these interactions might prove deleterious to cardiovascular function. Increased epicardial adipogenesis induces the secretion of pro-inflammatory adipokines which contribute to atrial and ventricular damage^7,12,19^. In the setting of COVID-19, EAT is postulated to play a role in generating the imbalance between the production of anti and pro-inflammatory adipokines, thus contributing to the enhanced cytokine storm and multisystem involvement documented in severe COVID-19^8^. In this study, EAT thickness was a significant predictor of adverse biochemical profiles, severe COVID-19 and increased mortality independent of BMI.

Using mediation analyses, EAT thickness was shown to play a significant role in mediating the risk of severe COVID-19 and mortality attributable to increasing BMI. Other authors have shown that increased VAT deposition has a greater predictive value for adverse COVID-19 outcomes and overall cardio-metabolic risk compared to BMI alone as a measure of whole-body adiposity. Adipose tissue dysfunction may be particularly relevant in younger cases and may display dysfunction based on specific anatomical locations, particularly when comparing subcutaneous to visceral adipose tissue distribution^20–23^. These results are consistent with previous findings in this population and further contribute to the characterization of the mechanisms underlying the risk attributable to aging and obesity in severe COVID-19 ^25–28^. Increased BMI has been shown to increase the risk of severe COVID-19; however, as BMI is an imperfect measure of body composition, this effect may be partly mediated by the relationship between BMI and EAT. Increased chronological age has similarly been shown to increase risk of severe COVID-19 and this effect may be partly mediated through the effect of age in increasing metabolically unfavorable fat depots, including VAT and EAT. Therefore, cases with low BMI but increased EAT were generally older; conversely, cases with increased BMI and increased EAT were as young as non-obese cases and cases without increased EAT. This interplay shows that EAT may increase risk in cases otherwise perceived as low risk, including lean and younger individuals.

Finally, increased EAT accumulation was shown to be associated with increased cardiac damage and an increased hypercoagulable state through its effect on fibrinogen levels, thus contributing to risk for COVID-19 severity and mortality. Increased expression of ACE II in whole-body and visceral adipose tissue in persons with obesity has been proposed as a potential mechanism for increasing the risk of COVID-19 by promoting viral spread and shedding and by amplifying the inflammatory responses ^24–26^. Increased risk of cardiovascular complications in COVID-19 have been associated with the pro-thrombotic and inflammatory adaptive responses to SARS-CoV-2 infection as well as direct damage to the myocardium^9,27,28^. Increased EAT thickness was associated with increased risk of in-hospital myocardial infarction and was associated with increased cardiac troponin levels, possibly indicating direct cardiovascular injury. Since EAT is only a measure of ectopic fat deposition around the myocardium, it can be considered complementary to BMI. Hence, it may be most useful to assess risk for potential cardiac involvement in COVID-19.

This work has certain strengths and limitations. The study analyses a large number of patients in whom EAT thickness was measured at admission and who were followed throughout hospitalization to record COVID-19 relevant outcomes. Furthermore, this work is novel as it assesses the different types of intrathoracic adipose tissue, including EAT, sub-thoracic and extra-pericardial in addition to whole-body adipose tissue measures such as BMI. Using mediation analyses, the degree of contribution of EAT in risk of severe disease and mortality was also quantified. Limitations include the use of EAT thickness instead of volume; this was because the CT scans on admission were simple scans in which volume could not be assessed. There were only a small number of recorded cardiovascular events, this means the full impact of EAT on COVID-19 cardiovascular complications could not be evaluated. The results only apply to moderate or severe infection as mild cases were not included due to the nature of case recruitment. The study was carried out at a single COVID-19 centre, and the findings may not be representative at a population level. Finally, since abdominal CT scans were not available for most patients, the contribution of thoracic vs. abdominal fat could not be assessed, an area which should be evaluated in future studies. In conclusion, EAT thickness was associated with increased risk for severe COVID-19 and mortality compared to other types of intrathoracic adipose tissue. EAT was also associated with increased cardiac troponin levels, suggesting it may play a role in the risk for cardiovascular complications in COVID-19. Furthermore, EAT appears to be a mediator of the effect of BMI on adverse disease outcomes. This work contributes to the body of evidence suggesting that EAT may be an important protagonist in adverse metabolic and inflammatory responses to SARS-CoV-2.

## Data Availability

DATA AVAILABILITY: Data is available from the corresponding author upon reasonable request. Code for reproducibility of results available at: https://github.com/oyaxbell/covid_metabolism

## CONFLICT OF INTEREST/FINANCIAL DISCLOSURE

Nothing to disclose.

## FUNDING

This research did not receive any specific grant from funding agencies in the public, commercial, or not-for-profit sectors.

## ACKNOWLEDGMENTS

All authors approved the submitted version. AVV, and NEAV are enrolled in the PECEM program at the Faculty of Medicine of UNAM; AVV and NEAV are supported by CONACyT.

## CONTRIBUTIONS

research idea and study design: RM, LMA, MRF; data acquisition: RM, NRM, BRE, IJA, JCVG, CIPC, PEVC, DHJ, CAAS; data analysis/interpretation: OYBC, RM, CAAS, NEAV, AVV; statistical analysis: OYBC; manuscript drafting: RM, OYBC, NEAV, AVV, PAV, DEL, CAAS; supervision or mentorship: RM, AGF, AGH, APLG, JSO, CAAS. Each author contributed important intellectual content during manuscript drafting or revision and accepts accountability for the overall work by ensuring that questions pertaining to the accuracy or integrity of any portion of the work are appropriately investigated and resolved.

## DATA AVAILABILITY

Data is available from the corresponding author upon reasonable request. Code for reproducibility of results available at: https://github.com/oyaxbell/covid_metabolism

## Notes

**CONFLICT OF INTERESTS:** Nothing to disclose.

### Competing Interest Statement

The authors have declared no competing interest.

### Author Declarations

All proceedings were approved by the research and ethics committee of the INCMNZ (Ref. 3383) and informed consent was waived due to the nature of the study.

### Summary of Updates

Paper with revisions after peer review. Extensive change of discussion and results section.

## REFERENCES

1 Huang Y, Lu Y, Huang Y-M, Wang M, Ling W, Sui Y et al. Obesity in patients with COVID-19: a systematic review and meta-analysis. Metabolism 2020; 113: 154378.

2 Vepa A, Bae JP, Ahmed F, Pareek M, Khunti K. COVID-19 and ethnicity: A novel pathophysiological role for inflammation. Diabetes Metab Syndr 2020; 14: 1043–1051.

3 Chiappetta S, Sharma AM, Bottino V, Stier C. COVID-19 and the role of chronic inflammation in patients with obesity. Int J Obes 2005 2020; 44: 1790–1792.

4 Korakas E, Ikonomidis I, Kousathana F, Balampanis K, Kountouri A, Raptis A et al. Obesity and COVID-19: immune and metabolic derangement as a possible link to adverse clinical outcomes. Am J Physiol Endocrinol Metab 2020; 319: E105–E109.

5 Bello-Chavolla OY, Antonio-Villa NE, Vargas-Vázquez A, Viveros-Ruiz TL, Almeda-Valdes P, Gomez-Velasco D et al. Metabolic Score for Visceral Fat (METS-VF), a novel estimator of intra-abdominal fat content and cardio-metabolic health. Clin Nutr Edinb Scotl 2020; 39: 1613–1621.

6 Unamuno X, Gómez-Ambrosi J, Rodríguez A, Becerril S, Frühbeck G, Catalán V. Adipokine dysregulation and adipose tissue inflammation in human obesity. Eur J Clin Invest 2018; 48: e12997.

7 Sato F, Maeda N, Yamada T, Namazui H, Fukuda S, Natsukawa T et al. Association of Epicardial, Visceral, and Subcutaneous Fat With Cardiometabolic Diseases. Circ J Off J Jpn Circ Soc 2018; 82: 502–508.

8 Malavazos AE, Goldberger JJ, Iacobellis G. Does epicardial fat contribute to COVID-19 myocardial inflammation? Eur Heart J 2020; 41: 2333.

9 Dhakal BP, Sweitzer NK, Indik JH, Acharya D, William P. SARS-CoV-2 Infection and Cardiovascular Disease: COVID-19 Heart. Heart Lung Circ 2020; 29: 973–987.

10 Driggin E, Madhavan MV, Bikdeli B, Chuich T, Laracy J, Biondi-Zoccai G et al. Cardiovascular Considerations for Patients, Health Care Workers, and Health Systems During the COVID-19 Pandemic. J Am Coll Cardiol 2020; 75: 2352–2371.

11 Knight SR, Ho A, Pius R, Buchan I, Carson G, Drake TM et al. Risk stratification of patients admitted to hospital with covid-19 using the ISARIC WHO Clinical Characterisation Protocol: development and validation of the 4C Mortality Score. BMJ 2020; 370: m3339.

12 Silaghi A, Piercecchi-Marti M-D, Grino M, Leonetti G, Alessi MC, Clement K et al. Epicardial adipose tissue extent: relationship with age, body fat distribution, and coronaropathy. Obes Silver Spring Md 2008; 16: 2424–2430.

13 Bertaso AG, Bertol D, Duncan BB, Foppa M. Epicardial fat: definition, measurements and systematic review of main outcomes. Arq Bras Cardiol 2013; 101: e18–28.

14 Mahabadi AA, Massaro JM, Rosito GA, Levy D, Murabito JM, Wolf PA et al. Association of pericardial fat, intrathoracic fat, and visceral abdominal fat with cardiovascular disease burden: the Framingham Heart Study. Eur Heart J 2009; 30: 850–856.

15 Márquez-Salinas A, Fermín-Martínez CA, Antonio-Villa NE, Vargas-Vázquez A, Guerra EC, Campos-Muñoz A et al. Adaptive metabolic and inflammatory responses identified using accelerated aging metrics are linked to adverse outcomes in severe SARS-CoV-2 infection. medRxiv 2020; : 2020.11.03.20225375.

16 Neeland IJ, Ross R, Després J-P, Matsuzawa Y, Yamashita S, Shai I et al. Visceral and ectopic fat, atherosclerosis, and cardiometabolic disease: a position statement. Lancet Diabetes Endocrinol 2019; 7: 715–725.

17 Stefan N. Causes, consequences, and treatment of metabolically unhealthy fat distribution. Lancet Diabetes Endocrinol 2020; 8: 616–627.

18 Smith U. Abdominal obesity: a marker of ectopic fat accumulation. J Clin Invest 2015; 125: 1790–1792.

19 Packer M. Epicardial Adipose Tissue May Mediate Deleterious Effects of Obesity and Inflammation on the Myocardium. J Am Coll Cardiol 2018; 71: 2360–2372.

20 Favre G, Legueult K, Pradier C, Raffaelli C, Ichai C, Iannelli A et al. Visceral fat is associated to the severity of COVID-19. Metabolism 2020; 115: 154440.

21 Watanabe M, Caruso D, Tuccinardi D, Risi R, Zerunian M, Polici M et al. Visceral fat shows the strongest association with the need of intensive care in patients with COVID-19. Metabolism 2020; 111: 154319.

22 Deng M, Qi Y, Deng L, Wang H, Xu Y, Li Z et al. Obesity as a Potential Predictor of Disease Severity in Young COVID-19 Patients: A Retrospective Study. Obes Silver Spring Md 2020; 28: 1815–1825.

23 Iacobellis G, Malavazos AE, Ferreira T. COVID-19 Rise in Younger Adults with Obesity: Visceral Adiposity Can Predict the Risk. Obes Silver Spring Md 2020; 28: 1795.

24 Iacobellis G, Secchi F, Capitanio G, Basilico S, Schiaffino S, Boveri S et al. Epicardial Fat Inflammation in Severe COVID-19. Obesity 2020; 28: 2260–2262.

25 Malavazos AE, Corsi Romanelli MM, Bandera F, Iacobellis G. Targeting the Adipose Tissue in COVID-19. Obes Silver Spring Md 2020; 28: 1178–1179.

26 Ryan PM, Caplice NM. Is Adipose Tissue a Reservoir for Viral Spread, Immune Activation, and Cytokine Amplification in Coronavirus Disease 2019? Obes Silver Spring Md 2020; 28: 1191–1194.

27 Long B, Brady WJ, Koyfman A, Gottlieb M. Cardiovascular complications in COVID-19. Am J Emerg Med 2020; 38: 1504–1507.

28 Levi M, Thachil J, Iba T, Levy JH. Coagulation abnormalities and thrombosis in patients with COVID-19. Lancet Haematol 2020; 7: e438–e440.

